# Accurate Hepatic Fat Fraction Quantification Across Body Sizes Using Photon-Counting CT

**DOI:** 10.64898/2026.07.28.26359152

**Authors:** Xinhua Li, Cindy Kallman, Di Zhang, Chao Guo, Yifang Zhou

**Author notes:** Corresponding author Yifang Zhou, Ph.D. Department of Imaging, S. Mark Taper Foundation Imaging Center, M335, Cedars-Sinai Medical Center, 8700 Beverly Blvd., Los Angeles, CA 90048, USA.

## Abstract

**Objective:** To identify common photon-counting CT (PCCT) virtual monochromatic imaging (VMI) settings for accurate hepatic fat fraction (FF) quantification across different body sizes, including large body habitus.

**Methods:** Six non-iodinated fat lesions (FF 5%–40%) were embedded in anthropomorphic liver phantoms representing medium-sized (25×32.5 cm^2^) and large (31×39 cm^2^) abdomens. Phantoms were scanned on a PCCT system (NAEOTOM Alpha) at 120 and 140 kV. CT numbers were measured in VMIs at 40–190 keV in 1-keV increments. Linear regression between the ground-truth FF and measured Hounsfield units (HU) was used to estimate FF. Common optimal VMI settings yielding the minimum relative root-mean-square error (rRMSE) in both phantoms were identified.

**Results:** A single VMI setting of 70 keV at 140 kV demonstrated the best overall performance across body sizes, with FF (%) = −0.689HU + 36.51 (R^2^ > 0.996), achieving rRMSE ≤ 3.4% and absolute RMSE ≤ 0.7% in both phantoms. Robust performance (rRMSE ≤ 5%) was consistently maintained across 69–71 keV using identical calibration parameters for both phantoms. These results represented a substantial improvement over previously reported dual-energy CT (DECT) performance, while enabling accurate quantification on PCCT at radiation doses approximately 40% lower than those used in prior DECT protocols.

**Conclusion:** PCCT enables accurate and robust hepatic fat fraction quantification independent of body size. A single protocol at 140 kV with VMIs of 69–71 keV consistently achieved low quantification errors, demonstrating strong potential for opportunistic liver fat assessment using PCCT, especially in obese patients.

## 1. Introduction

Metabolic dysfunction–associated steatotic liver disease (MASLD), formally known as non-alcoholic fatty liver disease (NAFLD), is defined by hepatic steatosis (≥5% of hepatocytes containing fat) and has been increasing in prevalence, estimated to affect more than 30% of adult population and 70-80% of individuals with obesity.[1–3] Severe obesity is associated with more advanced disease and worse clinical outcomes compared with lean MASLD.[4] MAFLD is understood to be one of the leading causes of cirrhosis and may progress to HCC. HCC may develop in MAFLD without cirrhosis. In addition to liver disease, MAFLD is associated with higher all-cause mortality.[5] Hepatic fat fraction (FF) serves as a quantitative imaging biomarker for noninvasive assessment of hepatic steatosis.[6] Accurate quantification of hepatic fat content in adult patients, especially in large patients, is essential for diagnosis, disease characterization and longitudinal assessment.[7,8] [9] While liver biopsy provides a histologic reference, its invasive nature and sampling variability motivate the development of reliable imaging-based alternatives.[7] Among available modalities including magnetic resonance imaging, computed tomography (CT) is particularly attractive for opportunistic fat assessment because abdominal CT is routinely performed for a wide range of clinical indications.[10–12]

Hepatic attenuation on CT is determined by the relative proportions of fat and normal liver parenchyma. Under ideal monochromatic conditions, Hounsfield units (HU) demonstrate a linear relationship with fat fraction.[12] However, conventional single-energy CT uses polychromatic x-ray spectra, and attenuation measurements are affected by beam hardening, scatter, spectral averaging, and image noise.[13,14] These effects can compromise the accuracy of FF quantification, particularly in patients with obesity and at lower fat fractions.[15–17]

Dual-energy CT (DECT) has been evaluated to mitigate these limitations by exploiting energy-dependent differences in x-ray attenuation. [18] However, current DECT technologies exhibit technical constraints that may affect quantitative performance in clinical practice. Virtual monochromatic images (VMIs) derived from DECT are not purely monochromatic and remain influenced by photon energy, spectral separation, patient size, and noise.[18,19] Dual-source DECT systems are further limited by restricted field-of-view (FOV) coverage of the secondary detector, which may result in truncation artifacts in large patients.[20] Fast kV-switching DECT offers broader FOV coverage but is constrained by reduced spectral separation and lower photon flux at low energies, particularly in patients with large body habitus.[14,20,21] Consequently, DECT-derived FF quantification can show systematic deviations from the ideal linear HU–FF relationship in obese individuals.[14]

Photon-counting CT (PCCT) enables direct detection and energy binning of individual photons, offering improved spectral resolution and material characterization compared with conventional energy-integrating detector systems. [20,22] Preliminary evidence suggests that PCCT may strengthen the linearity between CT attenuation and tissue composition, potentially enhancing the accuracy of hepatic fat quantification. [23–26][24,27] However, quantitative performance may still depend on the selected VMI energy level, as low- and high-energy bins are subject to reduced photon statistics, while low-energy images remain vulnerable to beam hardening and spectral effects, particularly in larger patients. The optimal VMI settings for accurate and robust fat fraction quantification across varying body sizes on PCCT have not been clearly established.

The purpose of this study was to evaluate the performance of PCCT virtual monochromatic images for hepatic fat fraction quantification and to identify the energy settings that provide the most accurate and linear results across different patient sizes, with specific emphasis on performance in large body habitus.

## 2. Materials and methods

### 2.1 Anthropomorphic liver phantoms and CT scans

This study utilized custom anthropomorphic liver phantoms (CIRS, Sun Nuclear, Norfolk, VA, USA) designed to closely mimic human anatomy and tissue attenuation. The phantoms included a realistically shaped liver, an embedded simulated spine, and surrounding soft tissue, divided into three segments along the craniocaudal axis (Figure 1). Six non-iodinated ellipsoidal lesions representing different degrees of hepatic steatosis were embedded within the liver, with predefined fat fractions of 5%, 10%, 15%, 20%, 30%, and 40%, and volumes ranging from 0.785 to 6.283 cm^3^. A medium-sized abdominal phantom (25 × 32.5 cm^2^) was used to represent typical adult body habitus. A large phantom (31 × 39 cm^2^) simulating obese patients was created by adding an airtight soft-tissue-equivalent ring around the medium phantom. Both phantoms had a craniocaudal length of 19 cm. The CT number specifications for the normal liver parenchyma and fat lesions at different energy levels are listed in Table 1.

**Table 1:** Ground-truth Hounsfield unit (HU) of simulated normal liver and fatty liver in the phantom.

| keV | HU -Normal Liver | HU- Fatty Liver (fat fraction) |  |  |  |  |  |
| --- | --- | --- | --- | --- | --- | --- | --- |
|  |  | 5% | 10% | 15% | 20% | 30% | 40% |
| 50 | 58.6 | 49.3 | 40.0 | 30.6 | 21.2 | 2.2 | -17.0 |
| 60 | 55.2 | 47.2 | 39.2 | 31.1 | 23.1 | 6.8 | -9.6 |
| 80 | 52.3 | 45.6 | 38.8 | 32.0 | 25.2 | 11.4 | -2.5 |
| 100 | 50.9 | 44.6 | 38.3 | 32.0 | 25.7 | 12.9 | 0.0 |
| 150 | 51.3 | 45.4 | 39.6 | 33.6 | 27.7 | 15.8 | 3.7 |

**Figure 1.**
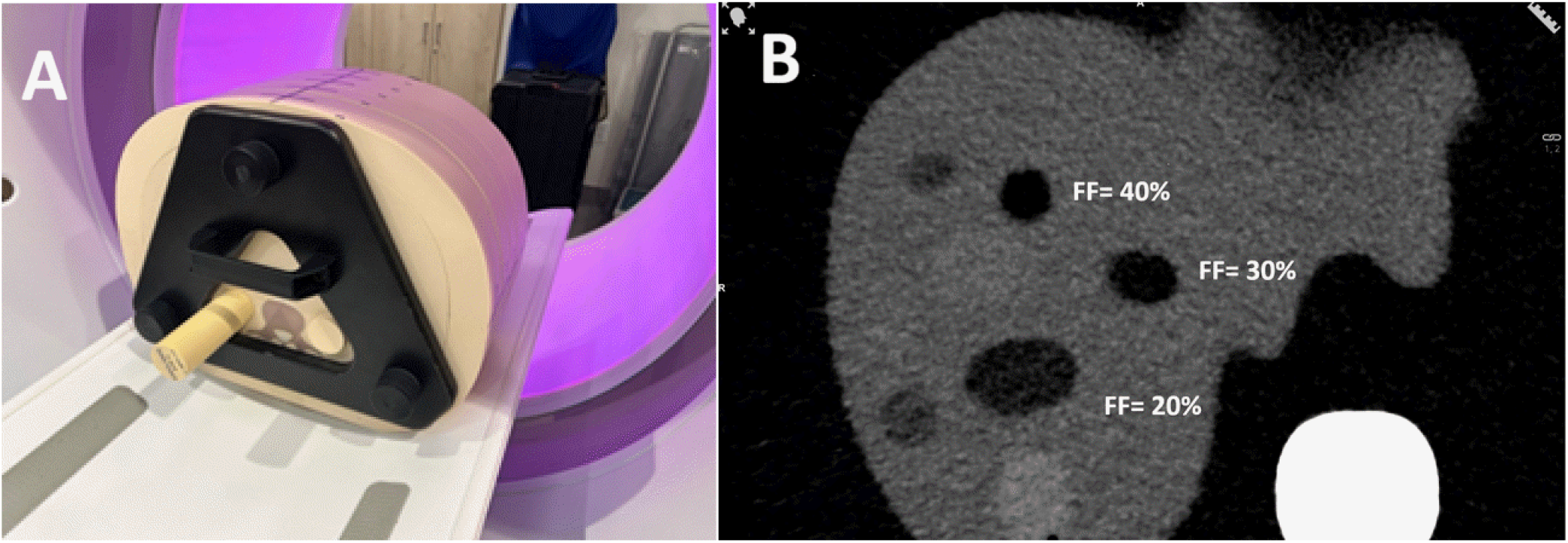
Anthropomorphic liver phantoms representing medium (25 × 32.5 × 19 cm^3^) and large (31 × 39 × 19 cm^3^) abdomens on the PCCT (A). The large phantom was configured by adding an airtight soft-tissue-equivalent ring around the medium-sized phantom. (B) Axial CT image at 70 keV showing three embedded fat lesions with fat fractions of 20%, 30%, and 40% within the liver.

Both phantoms were scanned on a NAEOTOM Alpha CT scanner (Siemens Healthineers, Erlangen, Germany) using a routine abdomen–pelvis protocol. Scans were performed in spectral mode (QuantumPlus) with four energy bin thresholds (20, 34, 62, and 75 keV), a scan FOV of 50.4 cm, beam collimation of 144 × 0.4 mm, and a helical pitch of 0.8, under automatic tube current modulation. Scans were acquired at 120 kV and 140 kV, with volume CT dose index (CTDI_vol_) of 12.8 mGy for the large phantom and 8.51 mGy for the medium-sized phantom. Each scan was repeated three times.

### 2.2 FF-HU relationship

Virtual monochromatic images were reconstructed at different energy levels using QR40f, QIR 3, and slice thickness 1.0 mm. The reconstruction FOV was 41.6 cm for the large phantom and 34.8 cm for the medium-sized phantom. Images were analyzed across 151 VMI energies (40–190 keV in 1-keV increments) on a Siemens Syngo.Via workstation (VB80F). At each VMI energy, CT numbers were measured in all six fat lesions using volumes of interest (VOIs) on images from three repeated scans. Measurements were performed with in-house developed software. All VOIs were placed entirely within the lesions, which had volumes of 0.14, 0.67, 0.76, 1.18, 1.20, and 2.15 cm^3^. Mean HU values from three repeat scans were used for linear regression against the known (ground-truth) fat fractions,[14]

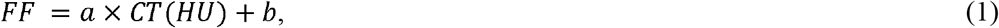

where *a* and *b* were regression coefficients.

For each phantom, the linear relationship between HU and FF was evaluated at 120 and 140 kV for every VMI energy by assessing coefficient of determination (R^2^). This resulted in a total of 604 fitted parameter pairs (a and b) (2 phantoms × 2 tube voltages × 151 energies). Data analysis was performed with statistical software R (version 4.4.2; The R Project for Statistical Computing, https://r-project.org).

### 2.3 Error analysis, body size specific optimal VMI energies

Across six lesions, the relative root-mean-square error (rRMSE) of the predicted fat fractions was calculated. This rRMSE was defined as the root-mean-square of the relative differences between the fat fractions derived from Equation 1 and the ground-truth fat fractions. The analysis was performed across 151 VMI energies for each phantom size and tube voltage, with the aim of identifying energies that minimized the rRMSE. These energies were denoted as body size-specific optimal VMI settings.

### 2.4 Body size independent optimal VMI energies

The optimal VMI settings identified for each phantom size were further evaluated across both phantoms and tube voltages to identify a common setting that achieved the minimum rRMSE independent of body size. This setting was denoted as the body size independent optimal VMI.

### 2.5 Lesion level errors

To provide a more comprehensive evaluation of FF quantification accuracy, errors were also calculated for each individual fat lesion at the identified optimal VMI energies. This analysis enabled a detailed assessment of the method’s performance across varying degrees of hepatic steatosis.

### 2.6 Comparison with prior dual energy results

Finally, the identified optimal VMI energies and corresponding error metrics were compared with previously reported dual-energy CT results.[14]

## 3. Results

### 3.1 FF-HU linearity assessment

The parameters of the linear regression varied with VMI energy, phantom size, and tube voltage, as summarized in Table 2. The negative slope became progressively steeper with increasing VMI energy. In contrast, the intercept remained relatively stable, varying only within a narrow range.

**Table 2:** Ranges of linear regression parameters (FF = a × CT [HU] + b) for both phantoms at 120 and 140 kV. The reported ranges reflect variations across the VMI energy range of 40–190 keV.

| | $a / b / R^2$ | |
| --- | --- | --- |
|  | Large | Medium-size |
| 120 kV | -0.38 to -0.99/35.6 to 36.6/0.957 to 0.996 | -0.45 to -0.88/33.8 to 33.7/0.973 to 0.999 |
| 140 kV | -0.43 to -0.84/32.1 to 40.5/0.971 to 0.996 | -0.40 to -0.92/34.6 to 37.8/0.972 to 0.999 |

The coefficient of determination (R^2^) varied with tube voltage, VMI energy, and phantom size, indicating differences in goodness of fit across acquisition conditions. Example FF–HU relationships corresponding to the lowest and highest R^2^ values are shown in Figures 2 and 3, respectively. The poorest fits were characterized by larger deviations at low fat fractions.

**Figure 2.**
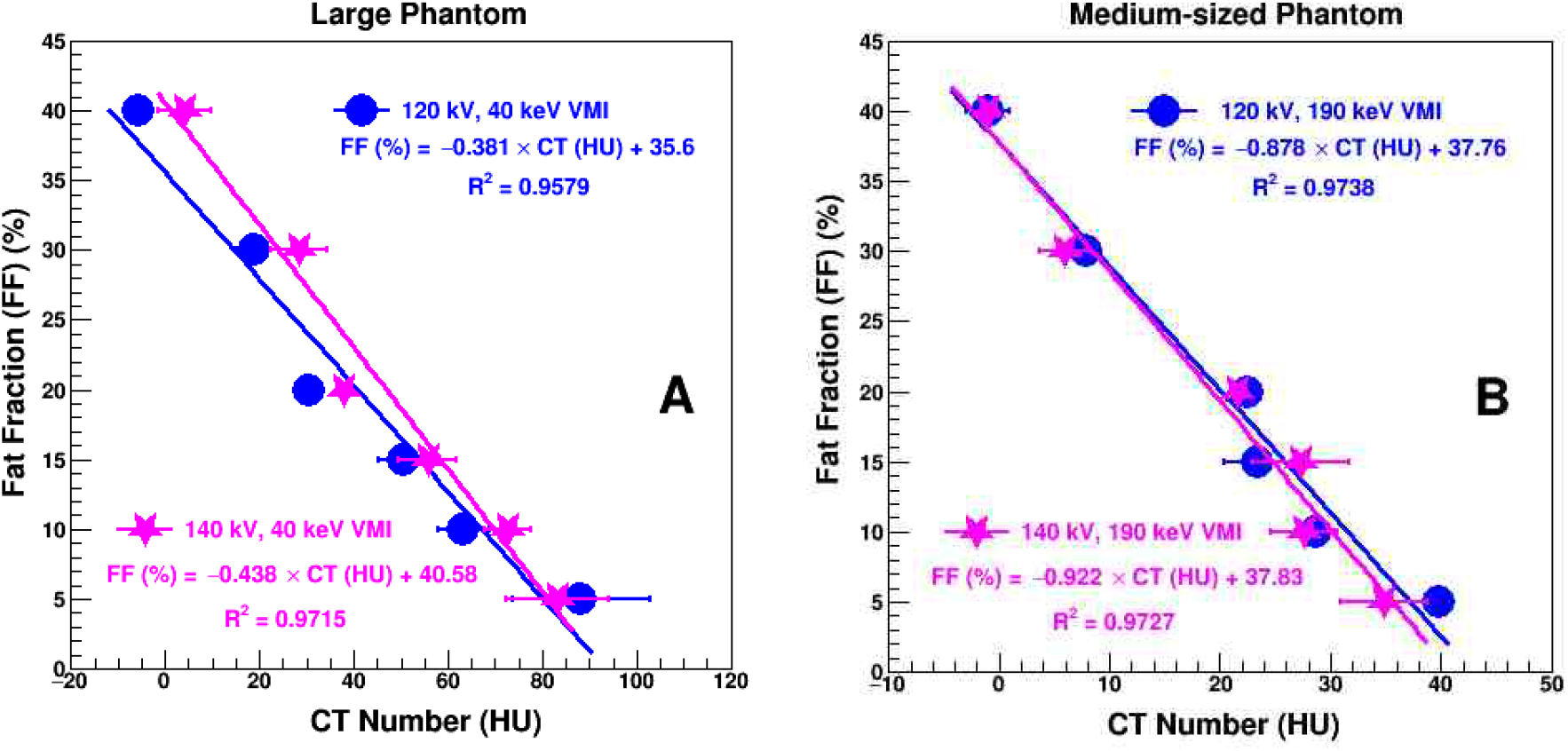
Illustration of fat fraction–CT number relationships at the VMI energy corresponding to the poorest goodness of fit. The panel shows the large phantom (A) and the medium-sized phantom (B), with linear regression parameters. Error bars show standard deviations of repeat scans.

**Figure 3.**
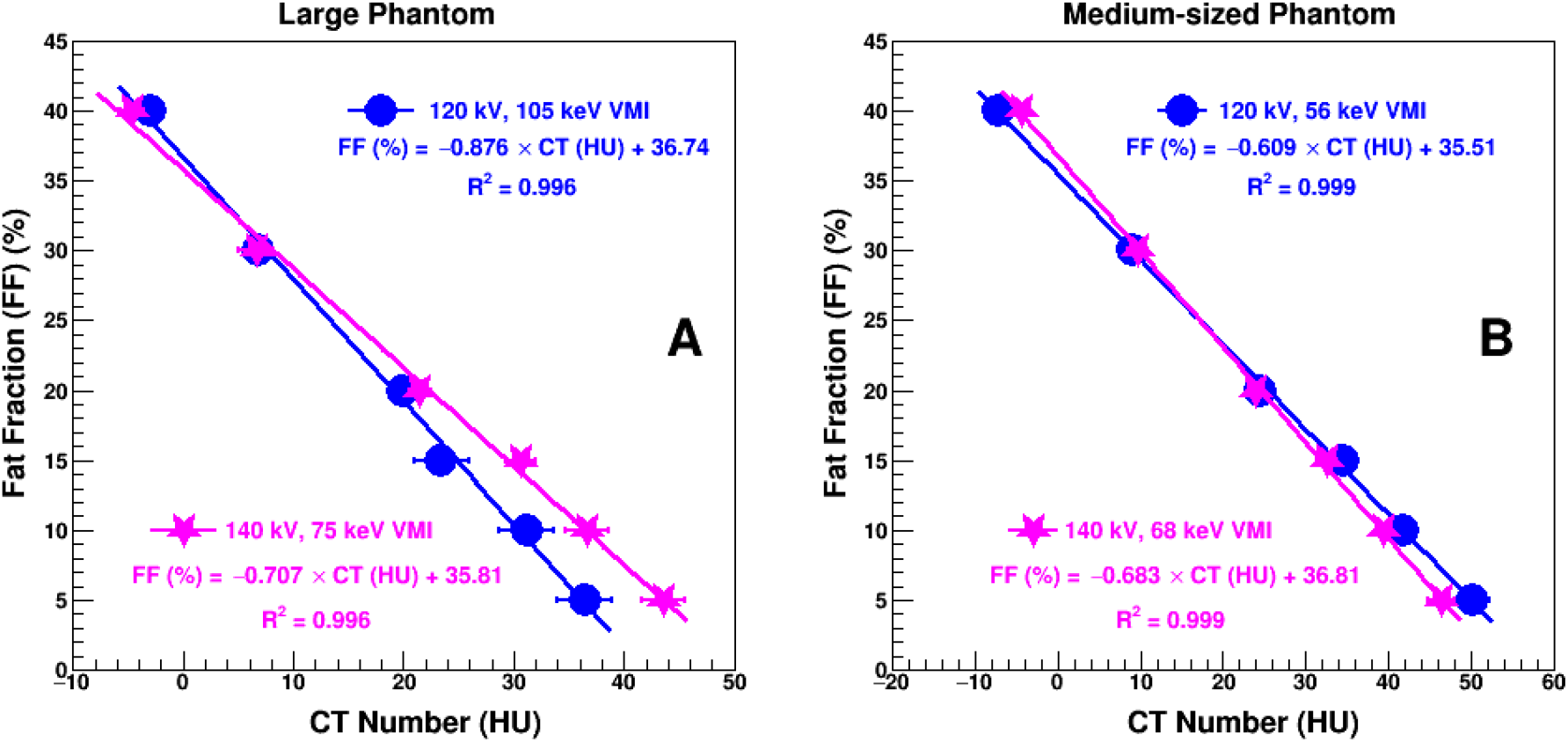
Illustration of fat fraction–CT number relationships at the VMI energy corresponding to the best goodness of fit. The panel shows the large phantom (A) and the medium-sized phantom (B), with linear regression parameters. Error bars show standard deviations of repeat scans.

### 3.2 Relative RMS errors of fat fraction estimates versus VMI energies

Figure 4 illustrates the relative root-mean-square error (rRMSE) of fat fraction estimates derived from the linear regression (Equation 1) across 151 VMI energies. The rRMSE varied with VMI energy, phantom size, and tube voltage. For the large phantom, rRMSE ranged from 4.59% to 27.0% at 120 kV and from 3.19% to 12.6% at 140 kV across all VMI energies. For the medium-sized phantom, rRMSE ranged from 1.88% to 22.0% at 120 kV and from 1.78% to 14.2% at 140 kV. Overall, 140 kV yielded narrower error ranges than 120 kV for both phantom sizes. For a predefined error threshold of ≤5%, 140 kV provided a broader VMI energy range than 120 kV. At 140 kV, the VMI energy ranges associated with rRMSE ≤5% were 56–85 keV and 59–98 keV for the medium-sized and large phantoms, respectively. In contrast, the corresponding ranges at 120 kV were 50–63 keV and 97–116 keV. The minimum rRMSE occurred at different VMI energies for each combination of phantom size and tube voltage.

**Figure 4.**
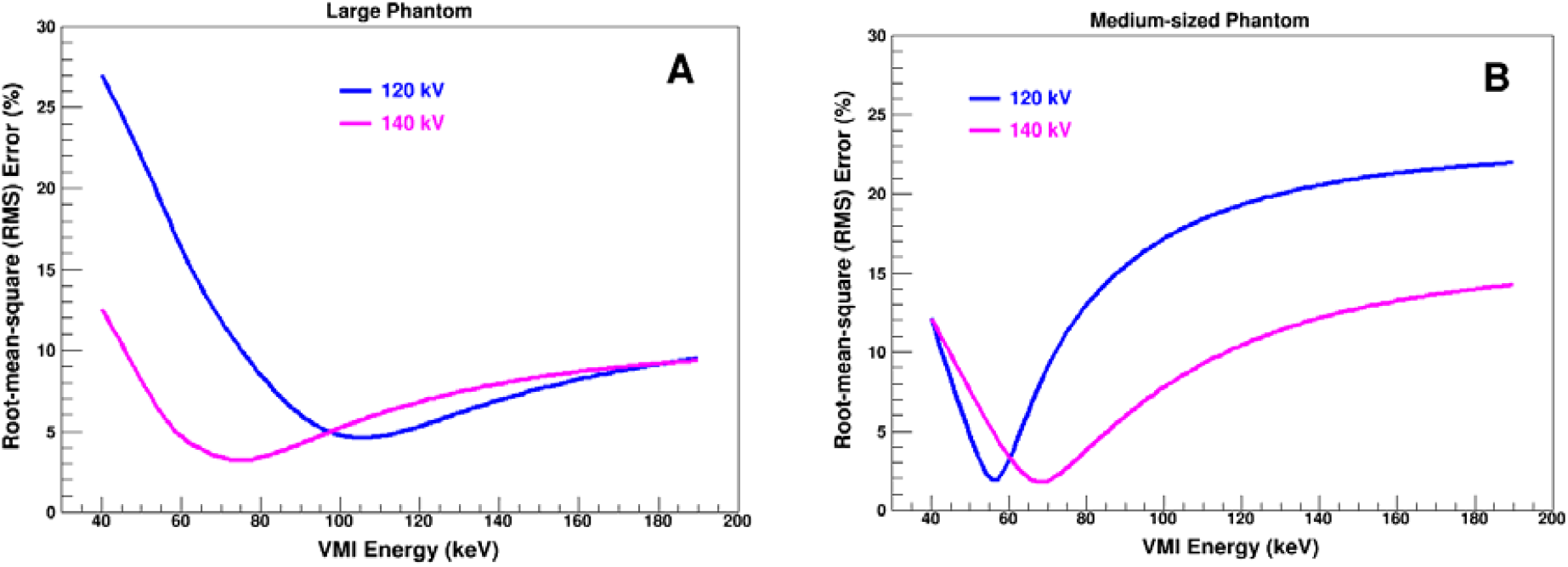
Relative root-mean-square (rRMS) errors of fat fraction estimates derived from linear regression (Equation 1) across virtual monoenergetic image (VMI) energies. The panel shows the large phantom (A) and the medium-sized phantom (B).

### 3.3 Body size specific optimal VMI energies

The VMI energy corresponding to the minimum rRMSE of fat fraction estimation was defined as the optimal VMI energy for each phantom size and tube voltage. The results are listed in Table 3.

**Table 3:** Optimal VMI energy and relative root-mean-square (rRMS) error from linear regression *FF* = *a* × *CT* (*HU*) + *b* for large and medium-sized phantom at 120 kV and 140 kV.

| Phantom | optimal keV/rRMS error (%) | | $a, b, R^2$ | |
| --- | --- | --- | --- | --- |
|  | 120 kV | 140 kV | 120 kV | 140 kV |
| Large | 105keV/4.6% | 75keV/3.2% | -0.876, 36.74, 0.996 | -0.707, 35.81, 0.996 |
| Medium-size | 56keV/1.9% | 68keV/1.8% | -0.609, 35.51, 0.999 | -0.683, 36.81, 0.999 |

### 3.4 Body size independent optimal VMI energies

As shown in Figure 4, the rRMSE versus keV curves for the large and medium-sized phantoms overlapped at 140 kV, while no such overlap was observed at 120 kV. At 140 kV, the minimum rRMSE occurred at 75 keV (3.2%) for the large phantom and at 68 keV (1.8%) for the medium-sized phantom. Given the broad low-error range around these energies and the lower rRMSE observed for the medium-sized phantom, an intermediate VMI energy was selected to minimize the increase in error for the large phantom while maintaining performance close to the optimum for the medium-sized phantom. A VMI energy of 70 keV was identified as the best compromise, corresponding to rRMSE values of 3.4% and 2.1% for the large and medium-sized phantoms, respectively. The corresponding slope and intercept were −0.689 and 36.51, respectively, derived from the weighted averages of the regression parameters obtained at 70 keV for both phantoms. When these parameters were applied to estimate fat fraction and compared with the known FF, robust performance (rRMSE ≤ 5%) was maintained over 69 –71 keV at 140 kV, as shown in Figure 5.

**Figure 5.**
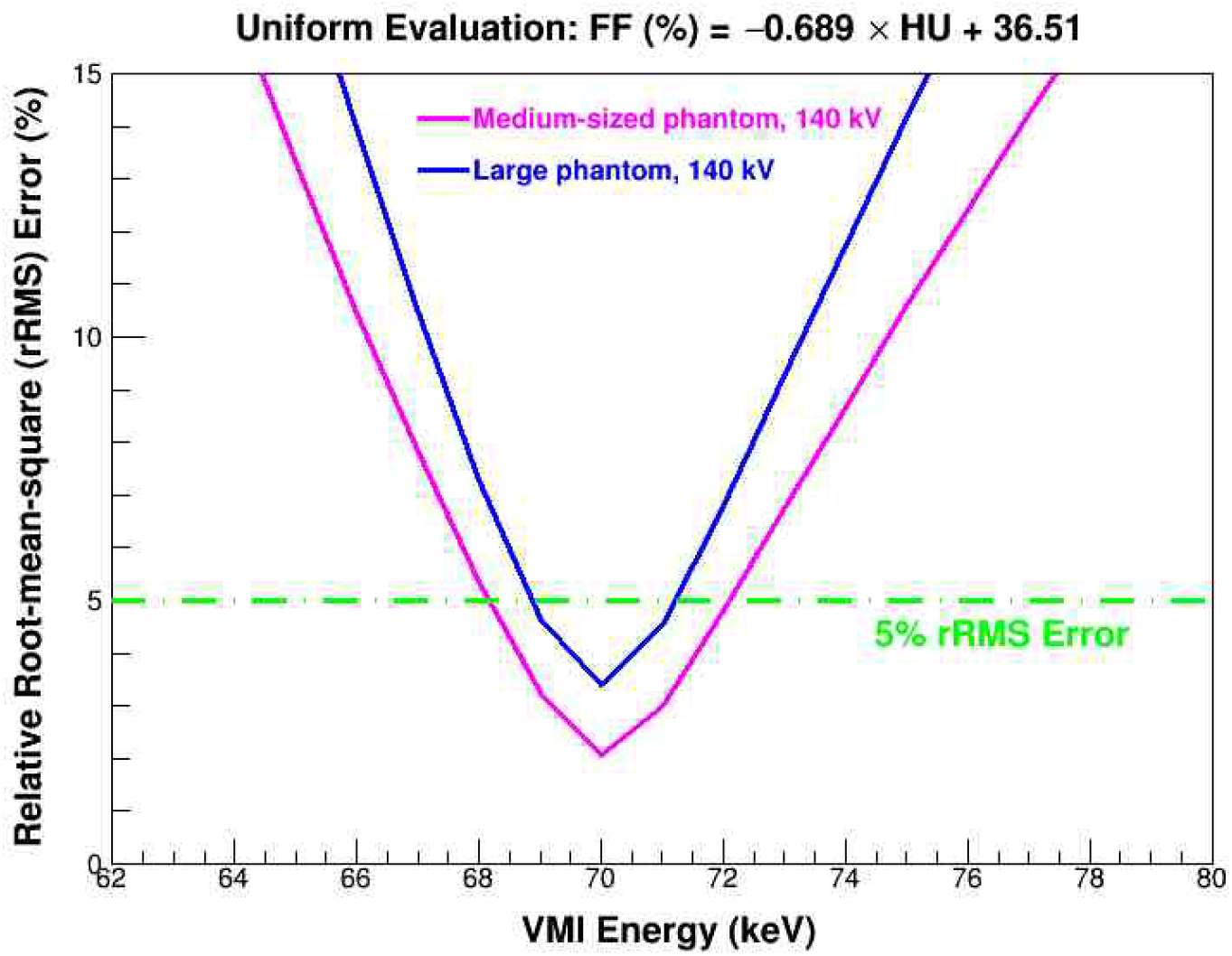
Relative root-mean-square errors (rRMSE) of fat fraction estimates using a slope of −0.689 and an intercept of 36.51 at 140 kV for the large and medium-sized phantoms.

**Figure 6.**
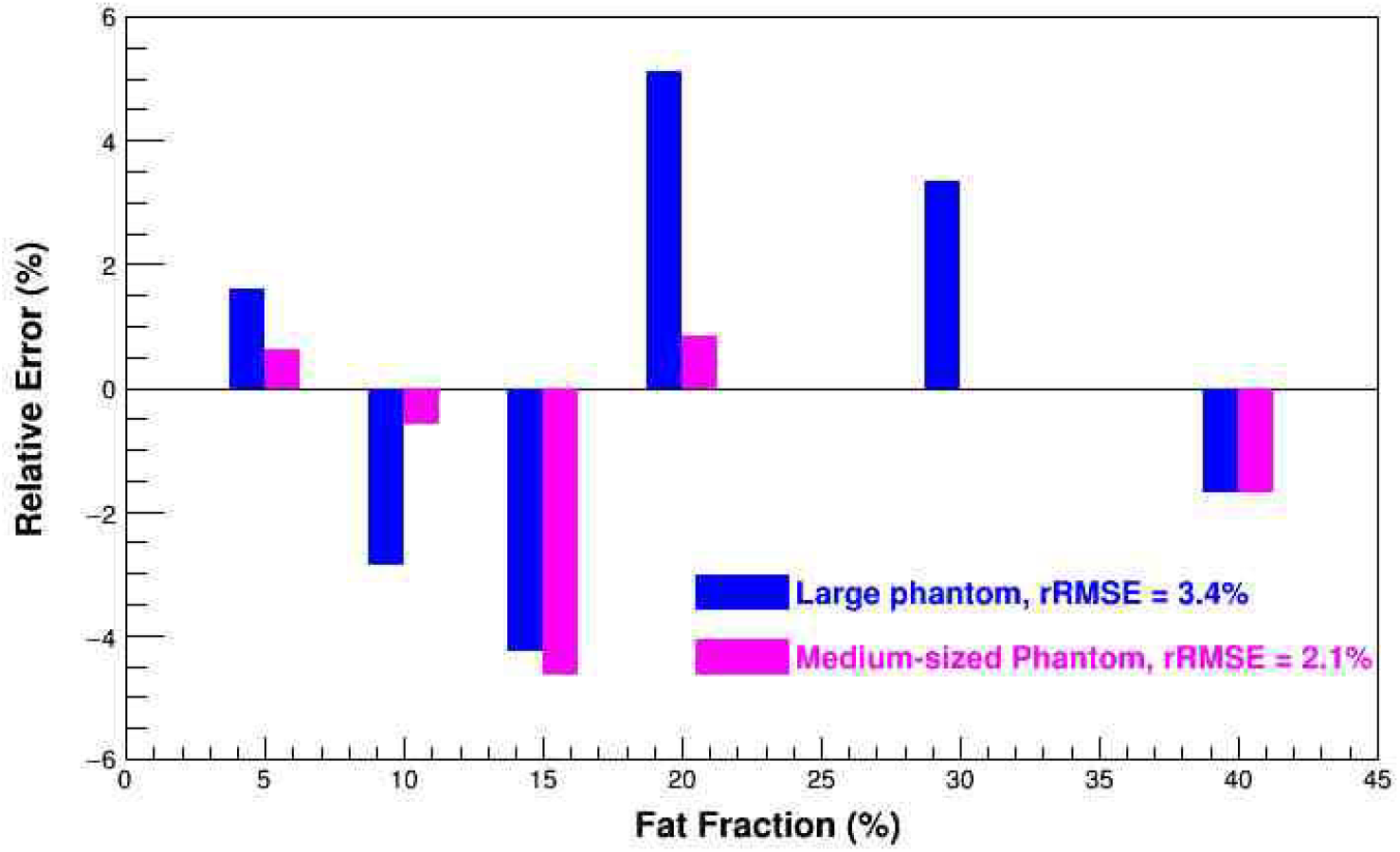
Liver fat fraction (FF) estimation errors for individual lesions at body size independent optimal VMI setting (70 keV/140 kV). The overall relative root-mean-square error (rRMSE) across six lesions is shown. The error for 30% FF in the medium-sized phantom was 0%.

### 3.5 Lesion level errors at body size independent optimal VMI energy

At the body size independent optimal VMI setting (70 keV/140 kV), relative errors ranged from −4.2% (FF,15%) to 5.1% (FF,20%) for the large phantom. For the medium-sized phantom, relative errors ranged from −4.6% (FF,15%) to 0.84% (FF,20%).

### 3.6 Comparison with dual energy VMI results

For comparison, corresponding DECT-derived VMI results have been reported previously [13]. Using fast kV-switching DECT, the minimum rRMSE for the large phantom was 15.3% at 140 keV, 80/140 kV (CTDI_vol_, 21 mGy), with a maximum lesion-level error of 27% at 10% fat fraction. For the medium-sized phantom, the minimum rRMSE was 4.8% at 100 keV, 80/140 kV (CTDI_vol_, 14 mGy), with a maximum lesion-level error of 10.6% at 10% fat fraction. No common optimal VMI setting was identified for the large and medium-sized phantoms. Using dual-source DECT, the minimum rRMSE for the medium-sized phantom was 2.4% at 120 keV, 80/150 kV (Sn) (CTDI_vol_,14 mGy), with a maximum lesion-level error of 3.9% at 30% fat fraction.

## Discussion

This study addressed the critical challenge of obesity-related inaccuracies in noninvasive hepatic fat quantification using CT. Although DECT enables FF assessment, its performance declines markedly in larger patients. PCCT holds promise for improved quantification. However, optimal VMI settings across body sizes have not been established. Using anatomically realistic liver phantoms with fat fractions of 5– 40% (mimicking early to established MASLD), we evaluated PCCT-derived VMIs (40–190 keV) in medium and large body habitus at 120 and 140 kV. In the large phantom (CTDI_vol_, 12.8 mGy), optimal VMIs achieved excellent accuracy with rRMSE of 3.2% (75 keV at 140 kV) to 4.6% (105 keV at 120 kV), substantially outperforming the previously reported DECT minimum rRMSE of 15.3% at 21 mGy. For the medium phantom (8.51 mGy), rRMSE values were 1.8% (68 keV at 140 kV) and 1.9% (56 keV at 120 kV), also superior to DECT (2.4% at 14 mGy).

More importantly, a single optimal VMI setting independent of the phantom size was identified to achieve 3.4% or lower rRMSE. The relative error at the low fat lesion (5%) was from 0.78% to 1.75%. An identical look-up-table from HU to fat fraction can be established based on the found equation: *FF(%)= -0*.*689HU+ 36*.*51 (R*^*2*^*> 0*.*996)*. No similar findings were reported in the prior DECT VMI study where the dose was 40% higher [13].

Figure 4 highlights that, independent of phantom size and tube voltage, rRMSE for FF quantification exhibits a characteristic U-shaped curve, decreasing initially with rising VMI energy before increasing at higher energies. This pattern enables identification of optimal VMI settings, with 70 keV at 140 kV providing accurate FF assessment across both medium and large body sizes. Accurate quantification (rRMSE ≤3.5%) is achievable over a relatively broad range at 140 kV (60–78 keV for medium and 69–82 keV for large phantoms). The largest errors occurred at 40 keV in the large phantom, likely due to increased beam hardening, scatter, and photon starvation, while the medium phantom showed highest errors at 190 keV, attributable to reduced high-energy photon counts. Optimal VMI energy shifted lower at 140 kV versus 120 kV in the large phantom, with the opposite trend observed in the medium phantom—differences possibly related to scanner-specific energy weighting. [28, 29][13]

In this study, we quantified hepatic fat using the linear relationship between HU and FF derived from VMIs on unenhanced scans. This approach enables simple, opportunistic liver fat assessment without the need for iodine contrast. Material decomposition was not used, as fat maps generated by the same commercial platform yielded inaccurate results, particularly at lower fat fractions. For instance, in the medium-sized phantom at 120 kV, measured FFs for true values of 5%, 10%, and 15% were 21.4%, 22.6%, and 26.2%, respectively. Even at higher fat fractions (20%, 30%, and 40%), the measured FFs only improved moderately to 28.2%, 38.3%, and 43.8%.

This study has limitations. Accuracy was evaluated using two phantom sizes, representing typical and large adult body habitus. The primary aim was to assess the feasibility of improving hepatic fat fraction quantification with PCCT in abdominal examinations, particularly in large patients, a population at high risk for MASLD. Although the phantom-based design allowed controlled evaluation across known fat fractions, further validation in clinical setting will be necessary for translation into routine practice.

With PCCT, the latest advancement in CT technology, accurate liver fat quantification remains challenging due to differences in patient body size, scanning techniques, and analysis methods. Although VMIs can be generated across a wide range of energy levels, the optimal setting often varies with body size, which can complicate everyday clinical use. This study offers a practical solution. Across both medium and large adult body sizes, we identified a single, reliable setting, 70 keV at 140 kV, combined with a simple conversion formula [FF(%) = −0.689 × HU + 36.51] that provides accurate liver fat ≤ quantification (rRMSE ≤ 3.4% and absolute RMSE ≤ 0.7%). This approach worked consistently well even with 40% lower radiation dose than DECT. Importantly, nearby settings (69–71 keV) produced similarly robust results with rRMSE ≤ 5%, offering flexibility in daily practice. The 70-keV VMI setting aligns with current emerging protocols for abdominal PCCT.[28] These findings support the use of routine, non-contrast abdominal CT, already performed for many clinical indications, for reliable detection and monitoring of MASLD, particularly in patients with obesity who are at highest risk.

## 5. Conclusion

A single VMI setting (70 keV at 140 kV) was identified across all subject sizes, achieving a relative RMS error of FF of ≤3.4% with photon counting CT and supporting emerging abdominal PCCT VMI protocols centered around approximately 70 keV. For individual subjects, FF estimates achieved relative errors of <5% and <2% in the large- and medium-sized phantoms, respectively, at a 40% lower dose than DECT.

As PCCT becomes more widely available, there is potential for significant clinical impact by enabling reliable, non-invasive assessment of liver fat for the diagnosis and monitoring of MAFLD. While the phantom-based design enabled controlled evaluation across known ground-truth fat fractions, further validation in clinical cohorts will be necessary before translation into routine patient care.

## Data Availability

All data produced in the present work are contained in the manuscript.

## Acknowledgement

The authors would like to thank Michael McNitt-Gray, Ph.D., for assistance with image acquisition. The authors also extend thanks to Alexander Scott, Ph.D., Nader Binesh, Ph.D., Emi Eastman M.S., and Christina Lee, B.S. for the helpful discussions.

